# Distinct T cell functional profiles in SARS-CoV-2 seropositive and seronegative children associated with endemic human coronavirus cross-reactivity

**DOI:** 10.1101/2023.05.16.23290059

**Authors:** Ntombi S. B. Benede, Marius B. Tincho, Avril Walters, Vennesa Subbiah, Amkele Ngomti, Richard Baguma, Claire Butters, Mathilda Mennen, Sango Skelem, Marguerite Adriaanse, Strauss van Graan, Sashkia R. Balla, Thandeka Moyo-Gwete, Penny L. Moore, Maresa Botha, Lesley Workman, Heather J. Zar, Ntobeko A. B. Ntusi, Liesl Zühlke, Kate Webb, Catherine Riou, Wendy A. Burgers, Roanne S. Keeton

## Abstract

SARS-CoV-2 infection in children typically results in asymptomatic or mild disease. There is a paucity of studies on antiviral immunity in African children. We investigated SARS-CoV-2-specific T cell responses in 71 unvaccinated asymptomatic South African children who were seropositive or seronegative for SARS-CoV-2. SARS-CoV-2-specific CD4+ T cell responses were detectable in 83% of seropositive and 60% of seronegative children. Although the magnitude of the CD4+ T cell response did not differ significantly between the two groups, their functional profiles were distinct, with SARS-CoV-2 seropositive children exhibiting a higher proportion of polyfunctional T cells compared to their seronegative counterparts. The frequency of SARS-CoV-2-specific CD4+ T cells in seronegative children was associated with the endemic human coronavirus (HCoV) HKU1 IgG response. Overall, the presence of SARS-CoV-2-responding T cells in seronegative children may result from cross-reactivity to endemic coronaviruses and could contribute to the relative protection from disease observed in SARS-CoV-2-infected children.

## INTRODUCTION

Severe Acute Respiratory Syndrome Coronavirus-2 (SARS-CoV-2) infection in children usually causes asymptomatic or mild illness, contrasting with the high rate of severe disease reported in older adults ^1,2^. As a result, global reports of coronavirus disease 2019 (COVID-19) cases among children and adolescents are underreported. The United Nations International Children’s Emergency Fund (UNICEF) estimates that 21% of all reported confirmed cases occur in individuals younger than 20 years ^3^.

In the USA, it has been documented that COVID-19-associated hospitalization rates among children less than 18 years were lower compared to those in older individuals ^4^. Indeed, the COVID-19-Associated Hospitalization Surveillance Network (COVID-NET) reported that children younger than 18 years accounted for only 4.2% of COVID-19-associated hospitalization and 0.2% of COVID-19-associated in-hospital deaths ^5^. In South Africa, during the first 2 years of the pandemic, 12.5% of confirmed cases and 0.7% of COVID-19 associated in-hospital deaths were in individuals younger than 19 years ^6^.

In contrast to the low COVID-19 severity in the majority of younger individuals, it is known that children are more susceptible than adults to other acute viral respiratory tract infections including respiratory syncytial virus (RSV), rhinovirus (RV), influenza virus and common circulating human coronaviruses (HCoV) ^7^. Several age-associated factors have been proposed to play a role in the reduction of severity to SARS-CoV-2 infection in children ^2,8^, including limited comorbidities ^9–11^, differences in the expression of SARS-CoV-2 viral entry factors ^12–14^, robust innate immune responses ^15–18^, humoral and cellular immunity ^19–21^ and pre-existing immunity against common cold circulating endemic HCoVs ^22–26^. Endemic HCoVs account for 15 to 30% of respiratory infections reported annually in children ^27,28^. These HCoVs belong to the alpha-coronavirus subfamily (HCoV-229E and HCoV-NL63) and the beta-coronavirus subfamily (HCoV-OC43 and HCoV-HKU1) and have seasonal infection peaks during the winter season and are responsible for high rates of infection among children ^29,30^.

It is well established that protective immune responses to SARS-CoV-2 encompass both an antibody as and T cell components ^31^. Several studies have reported robust and durable antibody and T cell responses against SARS-CoV-2, which are maintained up to 6-12 months following infection ^21,32–35^. However, data on T cell responses in children compared to adults are conflicting, with studies reporting lower SARS-CoV-2-specific T cell responses in children ^23,36,37^, no differences ^38^ or greater T cell responses in children ^22^. Many of these studies make use of ELISPOT assays which preclude analysis of T cell phenotypes or functional cytokine profiles.

In this study, we prospectively characterized specific T cell responses in SARS-CoV-2 seropositive and seronegative children during the COVID pandemic and determined the functional profiles of SARS-CoV-2-specific T cells, in the context of their pre-existing immunity against endemic beta-HCoVs. This study provides novel insights into cross-reactive immunity to SARS-CoV-2 in children.

## RESULTS

### Study cohort

To investigate SARS-CoV-2-specific immune responses, we measured immunoglobulin (Ig) G and T cell responses in 71 children recruited in Cape Town, Western Cape, South Africa. The participants are described in **Figure 1A**. The median age of the children was 7 years (interquartile range (IQR) 2.8-9 years) and 34% (24/71) were female. The children included in this study had not received any SARS-CoV-2 vaccine prior to recruitment and no PCR-confirmed infection data were available, although 58% showed SARS-CoV-2 seropositivity (defined as being positive for either anti-spike or anti-nucleocapsid IgG). Samples were collected between 1 February 2021 – 20 May 2021, after two infection waves in South Africa, that were dominated by the ancestral D614G strain followed by the Beta variant of concern ^39^.

**Figure 1:**
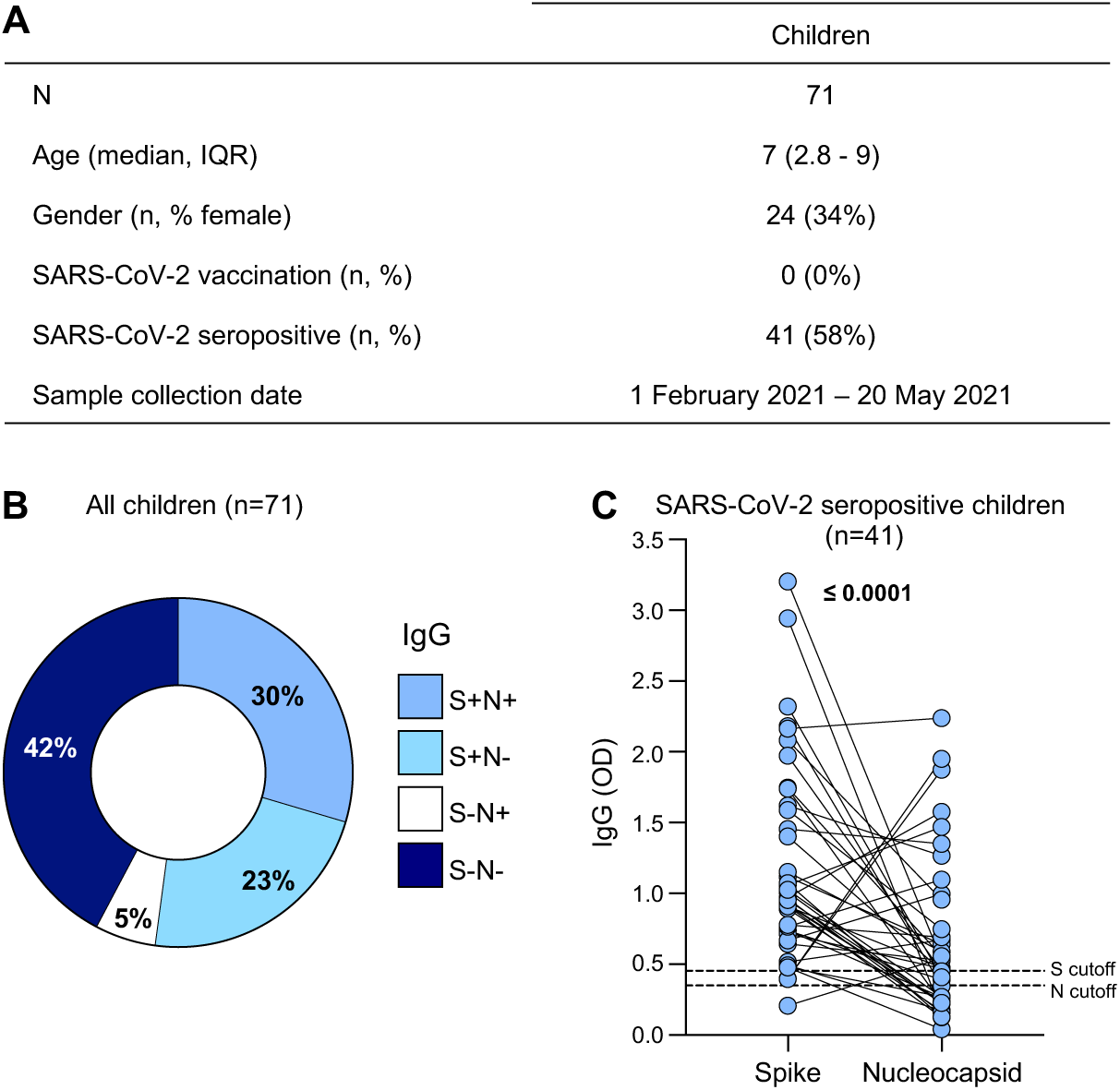
SARS-CoV-2-specific antibody responses in children. (A) The demographic characteristics of 71 unvaccinated children included in this study. Age, gender, SARS-CoV-2 vaccination status, SARS-CoV-2 serology and collection date are shown. (B) Proportion of children exhibiting antibody responses to SARS-CoV-2 spike (S) and nucleocapsid (N) proteins. (C) The magnitude of SARS-CoV-2 S and N IgG antibodies (OD_490nm_) measured by ELISA in seropositive children (n=41). The dotted lines indicate the cut-off for positivity which was calculated as the mean optical density of COVID-19 prepandemic control samples. Statistical analysis was performed using the Wilcoxon signed rank test; p values <0.05 were considered statistically significant and are bolded.

### SARS-CoV-2-specific antibody responses in children

To characterize the children serologically, we measured IgG responses against SARS-CoV-2 spike (S) and nucleocapsid (N) proteins using an indirect Enzyme-linked immunosorbent assay (ELISA) (**Figure 1B**). A large proportion (58%, 41/71) of unvaccinated children had detectable SARS-CoV-2-specific IgG against spike and/or nucleocapsid proteins and were classified as seropositive. Of the seropositive children, 30% (21/71) had spike– and nucleocapsid-specific IgG, 23% (16/71) had only spike-specific IgG and 5% (4/71) had only nucleocapsid-specific IgG only (**Figure 1B**). The remaining 42% (30/71) had undetectable SARS-CoV-2-spike and nucleocapsid-specific IgG and were classified as seronegative (**Figure 1B**). IgG responses to spike and nucleocapsid are shown for the 41 seropositive children in **Figure 1C**.

### SARS-CoV-2-specific T cell responses in children

The magnitude of SARS-CoV-2-specific T cell responses in children was quantified using a whole blood assay and intracellular cytokine staining followed by flow cytometry. SARS-CoV-2-specific CD4+ and CD8+ T cell responses were measured as total cytokine production of interferon-γ (IFN-γ), tumor necrosis factor-α (TNF-α) or interleukin-2 (IL-2) in response to a combined peptide pool covering SARS-CoV-2 spike (S), nucleocapsid (N) and membrane (M) proteins (**Figure 2A**).

**Figure 2:**
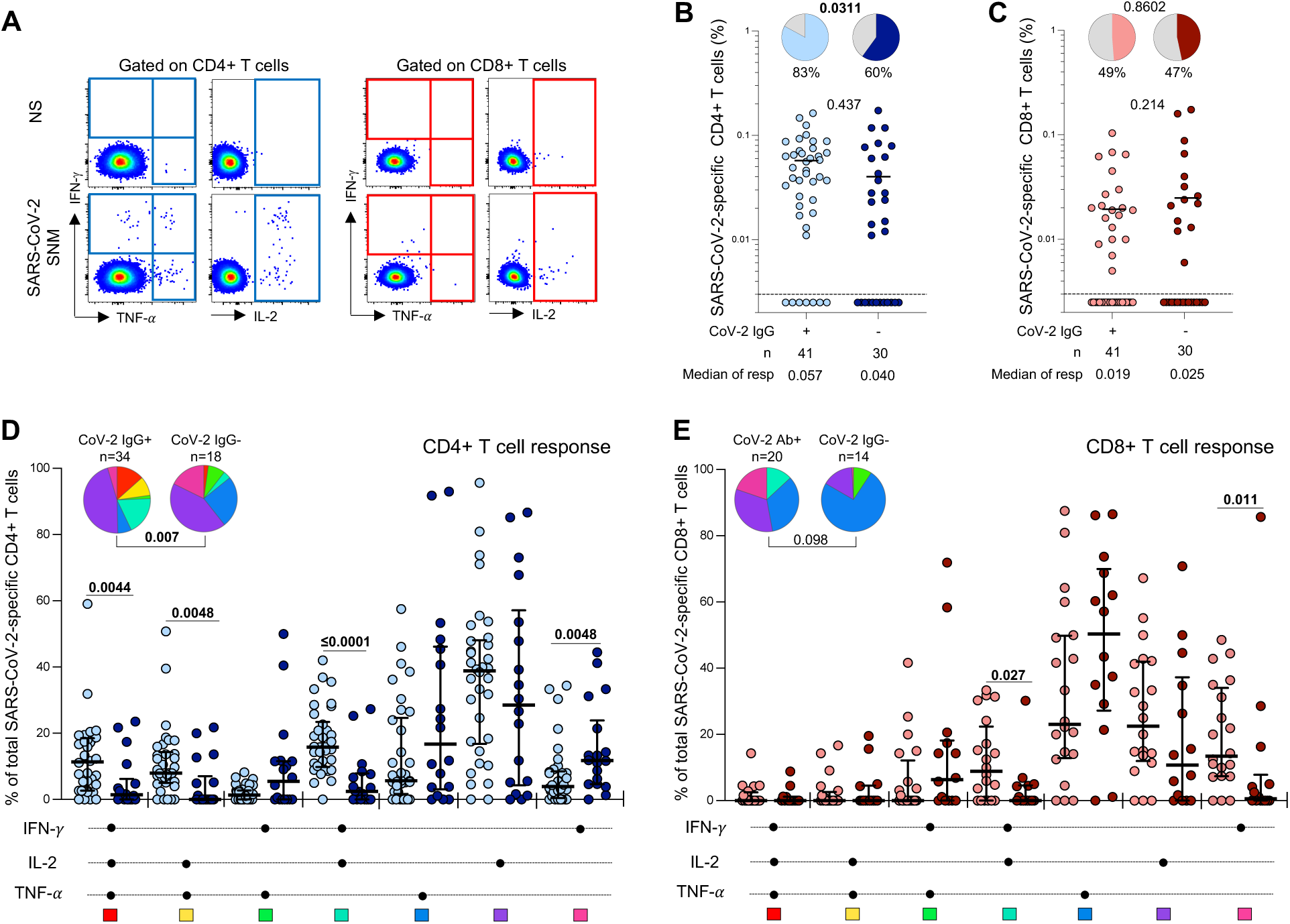
SARS-CoV-2-specific T cell responses in children. (A) Representative flow cytometry plots of SARS-CoV-2-specific interferon-γ (IFN-γ), tumor necrosis factor-α (TNF-α) and interleukin-2 (IL-2) cytokine production from CD4+ (left) and CD8+ (right) T cells in response to SARS-CoV-2 peptide stimulation. NS: no stimulation, SNM: combined peptide pool of SARS-CoV-2-spike, nucleocapsid and membrane proteins. (B) Frequency of SARS-CoV-2-specific CD4+ T cells producing any of the measured cytokines (IFN-γ, IL-2 or TNF-α). Children were grouped according to SARS-CoV-2 serostatus (light blue: n=41 SARS-CoV-2 seropositive; dark blue: n=30 SARS-CoV-2 seronegative). Bars represent the median of the responders, and median values are indicated. The pie charts represent the proportion of responders with detectable T cell response to SARS-CoV-2 SNM peptides. (C) Frequency of SARS-CoV-2-specific CD8+ T cells producing any of the measured cytokines (IFN-γ, IL-2 or TNF-α). Children were grouped according to SARS-CoV-2 serostatus (light red: n=41 SARS-CoV-2 seropositive children; dark red: n=30 SARS-CoV-2 seronegative children). Statistical comparisons in (B) and (C) were performed using the Mann-Whitney test between seropositive and seronegative children and the Chi-square test to compare the percentage of responders; p values <0.05 were considered statistically significant and are bolded. (D) Polyfunctional profile of SARS-CoV-2-specific CD4+ and (E) CD8+ T cells in seropositive and seronegative unvaccinated children. The x-axis illustrates each combination which is indicated with a black circle for the presence of IFN-γ, IL-2 and TNF-α. The medians and interquartile range are shown. Each response pattern (any possible combination of IFN-γ, IL-2 and TNF-α production) is color coded and summarized in the pie charts, with each pie slice representing the median contribution of each combination to the total SARS-CoV-2 responses. The permutation test was used to compare the statistical differences between the pie charts and the Mann Whitney Sum Test to compare response patterns between seropositive and seronegative children; p values <0.05 were considered statistically significant and are bolded.

We assessed the proportion of responders and magnitude of SARS-CoV-2-specific T cell responses according to the serostatus of the children (**Figure 2B and C**). As expected, 83% (34/41) of seropositive children had detectable SARS-CoV-2-specific CD4+ T cells. Interestingly, 60% (18/30) of seronegative children also had detectable SARS-CoV-2-specific CD4+ T cells, although this was a significantly lower proportion than observed in the seropositive group (p=0.0311). The magnitude of the SARS-CoV-2-specific CD4+ T cell response in seronegative responders was comparable to that observed in seropositive children (median of responders: 0.04% and 0.057% respectively; p=0.437, **Figure 2B**). Conversely, we did not observe significant differences in the proportion of SARS-CoV-2 CD8+ T cell responders (∼50%) or the frequency of SARS-CoV-2-specific CD8+ T cells between seropositive and seronegative children (median of responders: 0.019% and 0.025% respectively; p=0.214, **Figure 2C**).

Next, we evaluated the polyfunctional profile of SARS-CoV-2-responding CD4+ and CD8+ T cells based on the co-expression of IFN-γ, IL-2 or TNF-α (**Figure 2D, E**). The overall functional profile of SARS-CoV-2-specific CD4+ T cells in seropositive children was distinct from that of seronegative children (p=0.007, **Figure 2D**). SARS-CoV-2-specific CD4+ T cells in seropositive children were more polyfunctional, exhibiting a higher proportion of triple functional IFN-γ+IL-2+TNF-α+-producing CD4+ T cells (p=0.0044), and dual expressing cells (IL-2+TNF-α+, p=0.0048; IFN-γ+IL-2+, p≤0.0001) compared to the seronegative children. In contrast, seronegative children were characterized by an increased proportion of IFN-γ monofunctional CD4+ T cells (p=0.0048). Considering the CD8 compartment, we detected no significant differences in the overall functional profile of SARS-CoV-2-responding CD8+ T cells between the two groups (p=0.098), although seropositive children did have a larger proportion of CD8+ T cells producing both IFN-γ and IL-2 (p=0.027) and single IFN-γ (p=0.011) (**Figure 2E**).

Taken together, these data show that a significant proportion of seronegative children have detectable SARS-CoV-2-specific CD4+ and CD8+ T cells. Notably, these CD4+ T cells exhibit a distinct polyfunctional profile compared to those found in SARS-CoV-2-exposed children.

### Pre-existing immunity to endemic common circulating human coronaviruses

The presence of SARS-CoV-2-responding CD4+ T cells primarily exhibiting a monofunctional profile observed in seronegative children led us to hypothesize that the T cell response could be due to cross-reactivity resulting from prior infection with common circulating endemic HCoVs. Therefore, we measured endemic betacoronavirus HCoV-HKU1 and HCoV-OC43 spike IgG in SARS-CoV-2 seropositive and seronegative children. The magnitude of both HCoV-HKU1 (**Figure 3A**) and HCoV-OC43 (**Supplemental Figure 1A**) spike IgG was comparable in the seropositive and seronegative groups (median OD: 0.746 vs 0.420; p=0.153 for HCoV-HKU1 and 1.769 vs 1.281; p=0.07 for HCoV-OC43, respectively).

**Figure 3:**
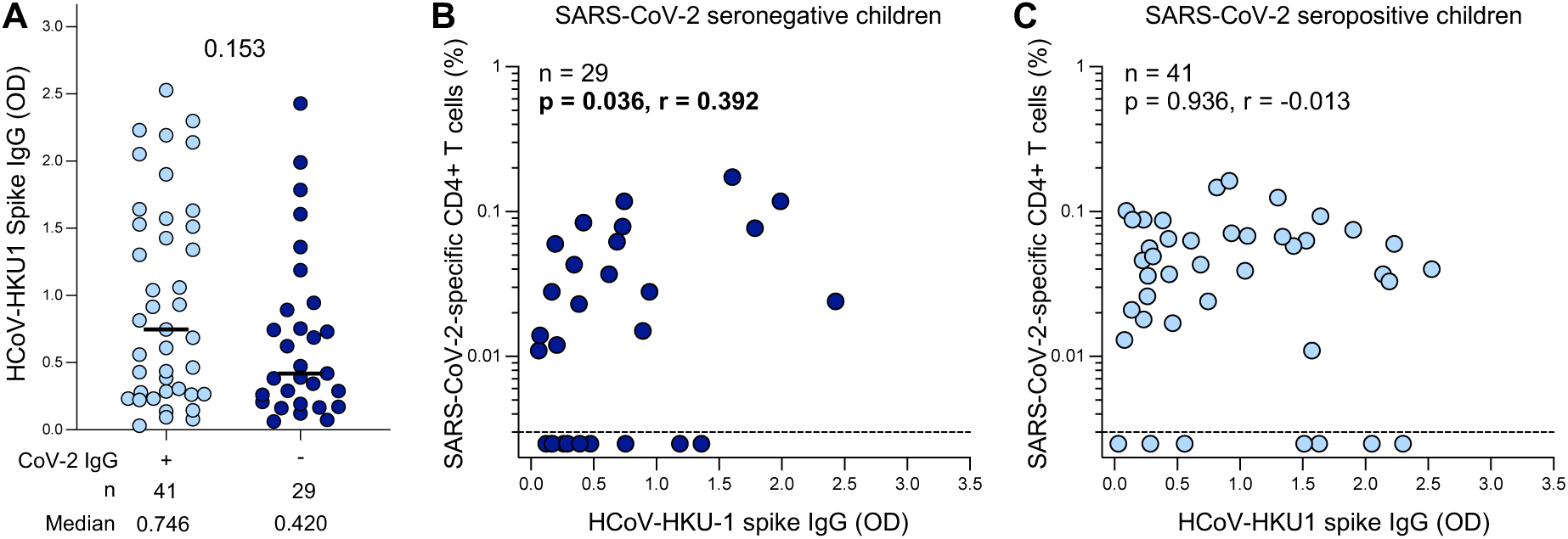
SARS-CoV-2 cross-reactivity to endemic beta-HCoV in children. (A) The magnitude of HCoV-HKU-1 spike IgG levels were measured by ELISA in SARS-CoV-2 seropositive (light blue; n=41) and seronegative (dark blue; n=29) children. Plasma sample was insufficient for one seronegative child therefore OD for HCoV-HKU1 was not measured for this participant. The bars represent the median values. A statistical comparison was performed using the Mann-Whitney test between seropositive and seronegative children; a p value <0.05 was considered statistically significant. (B) Correlation between the frequency of SARS-CoV-2-specific CD4+ T cells and HCoV-HKU-1-spike IgG levels in SARS-CoV-2 seronegative children (n=29). One participant had insufficient sample available to be included in this assay. (C) Correlation between the frequency of SARS-CoV-2-specific CD4+ T cells and HCoV-HKU-1-spike IgG levels in SARS-CoV-2 seropositive children (n=41). Statistical comparisons for (B) and (C) were performed using a two-tailed non-parametric Spearman rank tests; p values <0.05 were considered statistically significant and are bolded and correlation coefficients values are shown.

We found a significant correlation between the frequency of SARS-CoV-2-specific CD4+ T cells and HCoV-HKU1-spike IgG in seronegative children (p=0.036, r=0.392, **Figure 3B**), while no correlation was observed in seropositive children (p=0.936, r=-0.013, **Figure 3C**). For HCoV-OC43, no correlation was detected in either the seronegative (p=0.324, r=-0.187, **Supplemental Figure 1B**) or the seropositive group (p=0.171, r=-0.218, **Supplemental Figure 1C**). Overall, these results indicate SARS-CoV-2-reactive CD4+ T cell responses in seronegative children could be due in part to cross-reactive T cell immunity resulting from prior infection with common cold HCoV-HKU1.

### SARS-CoV-2-specific T cell responses in SARS-CoV-2 seropositive children compared to COVID-19 convalescent adults

Blood and plasma samples were collected from 30 COVID-19 convalescent healthcare workers (HCWs) participating in a longitudinal study at Groote Schuur Hospital (Cape Town, Western Cape, South Africa) (**Figure 4A**). All convalescent adults had a prior PCR-confirmed SARS-CoV-2 infection (median 224 days prior to sampling) and had not received a COVID-19 vaccine at the time of sampling. Samples were collected between 22 January 2021 – 23 February 2021.

**Figure 4:**
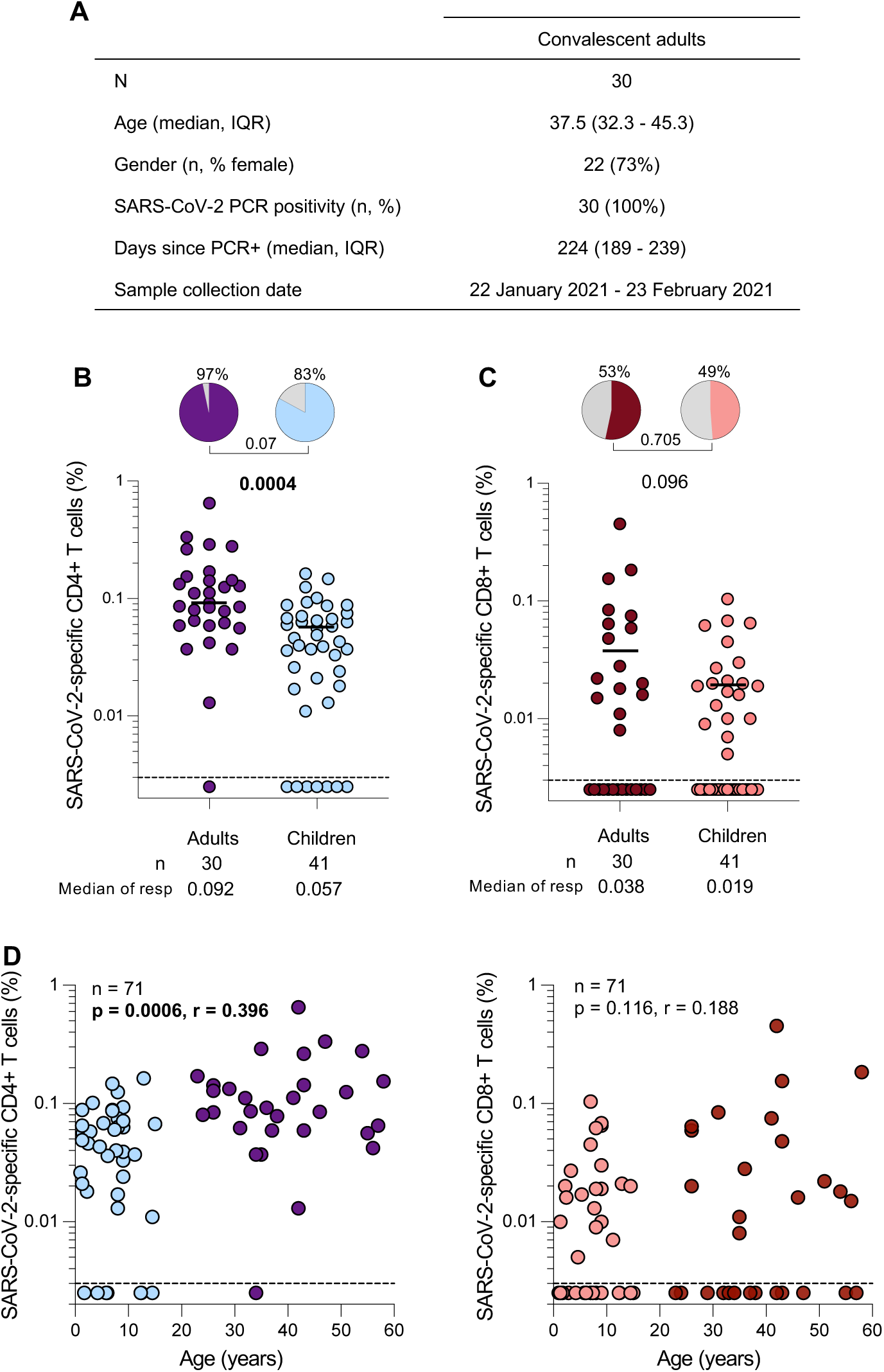
SARS-CoV-2-specific T cell responses in children compared to convalescent adults. (A) The demographic characteristics of 30 unvaccinated convalescent adults included in this study. Age, sex, SARS-CoV-2 PCR-positivity, days since PCR test and collection date are shown. (B) Frequency of SARS-CoV-2-specific CD4+ T cells producing any of the measured cytokines (IFN-γ, TNF-α, or IL-2) in SARS-CoV-2 convalescent HCW (purple; n=30) and seropositive children (light blue; n=41). The pie charts represent the proportion of responders with a detectable T cell response to SARS-CoV-2 SNM combined peptide pools. Bars represent median of the responders. (C) Frequency of SARS-CoV-2-specific CD8+ T cells producing any of the measured cytokines (IFN-γ, TNF-α, or IL-2) in SARS-CoV-2 convalescent HCW (dark red; n=30) and seropositive children (light red; n=41). The bars represent the median of the responders and median values are indicated. Statistical comparisons were performed using the Mann-Whitney test between seropositive children and adults and the Chi-square test was used to compare the percentage of responders; p values <0.05 were considered statistically significant and are bolded. (D) Correlations between SARS-CoV-2-specific CD4+ or CD8+ T cells and age in convalescent HCW (purple and dark red; n=30) and seropositive (light blue and light red; n=41) children. Statistical comparisons were performed using a two-tailed non-parametric Spearman rank tests; p values <0.05 were considered statistically significant and are bolded and correlation coefficients are shown.

A trend towards a higher proportion of SARS-CoV-2 CD4 responders was observed in convalescent adults (97%) compared to seropositive children (83%) (p=0.07, **Figure 4B**). Moreover, in SARS-CoV-2 responders, convalescent adults did have a significantly higher frequency of SARS-CoV-2-specific CD4+ T cells compared to seropositive children (median of responders: 0.092% vs 0.057%; p=0.0004, respectively). Unlike CD4+ T cell responses, there were no significant differences in either the proportion of CD8 responders (53% vs 49%, p=0.705) or the frequency of SARS-CoV-2-specific CD8+ T cells (median of responders: 0.038% vs 0.019%; p=0.096) between convalescent HCWs and seropositive children (**Figure 4C**). Furthermore, we observed a positive correlation between age and the frequency of SARS-CoV-2-specific CD4+ T cells (p=0.0006, r=0.396, **Figure 4D**). No correlation was observed with SARS-CoV-2-specific CD8+ T cells and age (p=0.116, r=0.188, **Figure 4E**). Overall, these results show that convalescent adults mount higher SARS-CoV-2-specific CD4 T cell responses compared to children, which could be attributed to age-related T cell development.

## DISCUSSION

T cells have been associated with protection from severe COVID-19 ^40^. It is therefore important to understand the nature of T cell responses targeting SARS-CoV-2 in children, who have been largely spared from severe COVID-19. In this study, comparing SARS-CoV-2-specific T cell responses in SARS-CoV-2 seropositive and seronegative children, a sizable proportion of seronegative children had SARS-CoV-2-reactive CD4+ and CD8+ T cells, but their CD4+ T cells exhibited a distinct functional profile compared to seropositive children. Importantly, in seronegative children, the frequency of SARS-CoV-2-reactive CD4+ T cells positively associated with HCoV-HKU1 spike-specific IgG antibodies. Additionally, we showed that convalescent adults had a higher magnitude of CD4+ T cell responses against SARS-CoV-2 compared to seropositive children, which associated positively with age.

Our data are in accordance with several studies showing that children develop robust humoral and cellular immunity to SARS-CoV-2 ^20,22,38,41,42^. In addition, SARS-CoV-2-specific T cell responses have been detected in 40-60% of SARS-CoV-2 unexposed individuals including children and adults ^22,23,43–48^ suggesting possible cross reactivity to HCoVs. One study demonstrated that the proportion of T cell responders was higher in children than adults ^22^. In contrast, a study by Tsang et al. found that SARS-CoV-2 uninfected children failed to mount detectable T cell responses ^41^. A further study showed that only a small proportion of seronegative children mounted a SARS-CoV-2-specific CD4+ T cell response (13%) compared to 60% in seropositive siblings, despite similar exposure in shared households ^42^. These two studies differ from our findings, where we showed that 60% of seronegative children had detectable SARS-CoV-2-specific T cell responses, consistent with Dowell et al ^22^. The discrepancies between the studies could be due to a difference in the seasonal prevalence of circulating HCoVs in each study setting ^29,30,49^, different T cell assays used to analyze T cell responses, and/or the cohort demographics.

Although the source of SARS-CoV-2 reactive T cells in seronegative children remains unclear, mounting evidence argues for cross-reactivity to endemic HCoVs. HCoVs (including beta HCoVs-HKU1 and OC43, and alpha HCoVs-NL63 and 229E) have partial sequence homology with SARS-CoV-2 ^44,50^. While cross reactive humoral responses have been shown not to underly this protection ^51^, studies have reported pre-existing T cell responses to endemic HCoVs with cross-reactivity to SARS-CoV-2 in unexposed adults ^47,52–57^ and children ^22,24,58,59^. These cellular responses were targeted mainly towards the S2 subunit which is highly conserved among coronaviruses ^22,25^. In our study, we showed a positive correlation between the frequency of SARS-CoV-2 CD4+ T cells and the magnitude of HKU1 spike-specific IgG in SARS-CoV-2 seronegative children. Our findings are supported by similar findings showing that 58% (7/12) seronegative children had binding antibodies against alpha and/or beta HCoV spike proteins, which they concluded were likely due to recent HCoV infection ^22^.

Pre-existing immunity from endemic HCoVs infection or exposure has been shown to be associated with a protective effect against COVID-19 disease in adults ^53,56,57,60,61^. Sagar et al. showed that individuals with a documented recent or ongoing HCoV infection and SARS-CoV-2 infection had fewer COVID-19-associated complications when hospitalized, compared to individuals without HCoV infection ^60^. Previously, the clinical relevance of pre-existing immunity and cross-reactive cellular responses in providing protection from infection in children was unclear. Recently however, Dowell et al. demonstrated that as for SARS-CoV-2 seropositive children, SARS-CoV-2-reactive T cell responses in SARS-CoV-2 seronegative children were protective against Omicron infection, suggesting a protective role in SARS-CoV-2 unexposed individuals ^62^.

Polyfunctional T cells have been reported to be associated with protection against a number of viral diseases ^63–66^. These include protection against cytomegalovirus (CMV) infection after lung transplantation ^64^, improved viral control of hepatitis C virus (HCV) ^65^ and detection during human immunodeficiency virus (HIV) infection in long term non-progressors ^66^. We found that seropositive children had a more polyfunctional profile of SARS-CoV-2-specific CD4+ T cells while CD4 responses in seronegative children exhibited a predominantly monofunctional profile. The SARS-CoV-2-specific polyfunctional Th1 CD4 response (characterized by co-expression of IFN-γ, TNF-α and/or IL-2), as seen in seropositive children, may be necessary for effective viral control and has been documented in COVID-19 convalescent adults ^67,68^.

The distinct functional profiles reported in our findings could be a result of differing degrees memory cell T cell differentiation or low SARS-CoV-2 peptide binding affinity with HCoV-specific T cells resulting in low T cell activation. The affinity between MHC molecules presenting peptide and the T cell receptor (TCR) plays a role in antigen recognition. It has been proposed that T cells with high affinity TCRs have greater effector functions and therefore an increased polyfunctional profile compared to low affinity TCRs ^69,70^. TCR cross-recognition of peptides on MHCs that are not structurally identical results in lower binding affinity which decreases T cell polyfunctionality ^70,71^. It is therefore plausible that the monofunctional profile of SARS-CoV-2 CD4+ T cell responses in seronegative children observed in our study may be mediated by cross-recognition of pre-existing T cell immunity to HCoV-HKU1, where partial sequence homology between HCoV-HKU1 and SARS-CoV-2 results in low peptide binding affinity and a different functional T cell profile ^44,55,69,71,72^. It is noteworthy to report that we did not find any association with the other beta HCoV-OC43 spike-specific IgG and SARS-CoV-2-specific T cells in seronegative children, which could be related to differences in seasonal prevalence of types of HCoV infections.

Numerous studies have now compared the magnitude of the SARS-CoV-2 specific T cell response in children to those detectable in adults ^22,23,36–38,41^. Initially it was thought that children may have a higher magnitude of T cell responses given their relative resistance to severe disease and the link between T cell responses and protection from severe disease ^8,73^. However, children were shown to have lower T cell responses than adults in most studies, including the current study ^23,36,37^. A plausible explanation is that adults have a mature immune system with more differentiated memory T cell subsets endowed with increased cytokine capacity, whereas children have an immature immune system, with many more naïve T cells which have reduced cytokine producing capacity, are enriched for monofunctional responses and have increased antigen dependence ^74^. Despite lower circulating SARS-CoV-2-specific T cells, children and adults may also have different responses in the respiratory tract. There is evidence in adults demonstrating the presence of SARS-CoV-2-specific T cell responses in the nasal mucosa after infection and vaccination ^75,76^. However comparative studies have yet to be performed in children. Additionally, several studies have proposed that one potential contributing factor to age-related COVID-19 clinical outcome are the robust innate immune responses observed in children, which may contribute to early control of viral replication ^15–17,77^. Recent studies have shown that, compared to adults, SARS-CoV-2 seronegative children exhibited an increased number of innate immune cells with pre-activated signatures leading to early production of interferon-mediated antiviral effects in the upper respiratory tract ^18,19^.

In conclusion, our study shows that a robust SARS-CoV-2-specific T cell response is observed in children, including in those with no evidence of prior SARS-CoV-2 infection. We demonstrate that the magnitude of SARS-CoV-2-reactive CD4+ T cells in seronegative children correlates with HCoV-HKU1 exposure. This, together with a distinct functional profile of SARS-CoV-2-specific responding CD4+ T cells observed between seropositive and seronegative children provides further evidence for pre-existing T cell responses cross-reactive to SARS-CoV-2.

### Limitations of the study

This study relied on serology to determine SARS-CoV-2 exposure. With no PCR confirmation of SARS-CoV-2 infection, the exact time of infection of the children could not be determined. Additionally, our study investigated T cell responses against SARS-CoV-2 spike, nucleocapsid and membrane proteins, and not the non-structural viral proteins that can also serve as targets for the T cell response, thus not capturing the full extent of T cell reactivity in infected participants. Furthermore, we did not address durability of the T cell responses in children due to the cross-sectional design of the study.

## **□**METHODS

### Lead contact

Further information and requests for resources and reagents should be directed to and will be fulfilled by the lead contact: Roanne S. Keeton.

### Materials availability

Materials will be made available by request to Roanne S. Keeton.

### Data and code availability

The published article includes all data generated or analyzed during this study, and summarized in the accompanying tables, figures, and supplemental materials.

## EXPERIMENTAL MODEL AND SUBJECT DETAILS

### Study participants

Paediatric participants (n=71) were recruited from two cohorts in the Western Cape, South Africa. The first cohort enrolled 50 children from the Red Cross War Memorial Children’s Hospital (Cape Town, Western Cape, South Africa). In this cohort 50/71 children were hospitalized for non-COVID-19-related elective procedures. A further 21 children were enrolled from the Drakenstein Child Health Study (DCHS) (Cape Winelands Western Cape, South Africa), a birth cohort study ^78^. The participants from this study were recruited between 1 February 2021 and 20 May 2021, after the first and the second infection waves with Ancestral strain and Beta variant in South Africa ^79^. Parents or legal guardians provided written informed consent for all paediatric participants. For the DCHS longitudinal cohort, this was renewed annually. The study was approved by the University of Cape Town Human Research Ethics Committee (HREC 599/2020 for the RC cohort and HREC 401/2009 for the DCHS cohort).

COVID-19 convalescent unvaccinated adults from a longitudinal study of healthcare workers (HCW) enrolled from Groote Schuur Hospital (Cape Town, Western Cape, South Africa) were included in the study (n=30). HCW in this cohort were recruited between July 2020 and January 2021 and were selected for inclusion based on a prior PCR-confirmed SARS-CoV-2 infection at least 3 months earlier. All participants were asymptomatic or had mild symptoms and did not require hospitalization for COVID-19 and were symptom-free at the time of sampling. Written informed consent was obtained from all participants and the study was approved by the University of Cape Town Human Research Ethics Committee (HREC 190/2020 and 209/2020).

## METHODS DETAILS

### SARS-CoV-2 and HCoV antigens

For serology assays, recombinant SARS-CoV-2 spike S1 (Cape Bio Pharms), and SARS-CoV-2 nucleocapsid (BioTech Africa) proteins were used for this study.

HKU1 and OC43 spike proteins were expressed in Human Embryonic Kidney (HEK) 293F suspension cells by transfecting the cells with the spike plasmid. After 6 days, proteins were purified using a nickel resin followed by size-exclusion chromatography. Relevant fractions were collected and frozen at –80°C until use.

SARS-CoV-2 peptides used for T cell assays included a commercially available peptide pool (15mers peptides with 11 amino acid overlap) covering the immunodominant regions of SARS-CoV-2 spike protein (PepTivator SARS-CoV-2 Prot_S, Miltenyi Biotech) based on the Wuhan-1 strain. The Spike peptide pool was prepared by resuspending in distilled water and used at a final concentration of 1 μg/ml. SARS-CoV-2 (Wuhan-1) nucleocapsid and membrane peptides (17mers with 10 amino acid overlap spanning the full proteins) were obtained from BEI Resources and were prepared by resuspending in dimethyl sulfoxide (DMSO, Sigma) and used at a concentration of 1 μg/ml.

### Enzyme-linked immunosorbent assay (ELISA)

SARS-CoV-2-specific enzyme-linked immunosorbent assay (ELISA) was performed to characterize the serostatus of participants, as previously described ^80^. Two ug/ml of spike protein was used to coat 96-well high-binding plates and incubated overnight at 4 °C. The plates were incubated in a blocking buffer consisting of 1% casein, 0.05% Tween 20, 1x Phosphate-Buffered Saline (PBS) for SARS-CoV-2 or 1x PBS, 5% skimmed milk powder, 0.05% Tween 20 for HCoVs. Plasma samples were diluted to 1:50 for SARS-CoV-2 or 1:100 for HCoVs in the respective blocking buffer. For the SARS-CoV-2 ELISA, secondary antibody was diluted to 1:5000 in dilution buffer and added to the plates followed by SigmaFast O-phenylenediamine dihydrochloride (OPD) substrate. For the HCoV ELISAs, secondary antibody was diluted to 1:3000 in blocking buffer and added to the plates followed by TMB substrate (Thermofisher Scientific). Upon stopping the reaction with 1-3 M sulfuric acid, absorbance was measured at a wavelength of 490 nm for SARS-CoV-2 or 450 nm for HCoVs. A cut-off for positivity was set at two standard deviations (SD) above the mean optical density (OD) of prepandemic samples for SARS-CoV-2 ELISAs

### Whole blood-based T cell assay

Blood was collected in sodium heparin tubes and processed within 4-6 hours of collection. The whole blood assay sample processing used for this study was adapted from a whole blood intracellular cytokine detection assay designed to detect SARS-CoV-2 specific T cells in adults ^81,82^. Briefly, 500 μl of blood was stimulated for 24 hours at 37°C with a combined pool of SARS-CoV-2 peptides including S, N and M, all at 1 µg/mL in the presence of costimulatory antibodies against CD28 (clone 28.2) and CD49d (clone L25) (1 μg/mL each; BD Biosciences) and Brefeldin A (10 μg/mL, Sigma-Aldrich). Unstimulated blood was incubated with costimulatory antibodies, Brefeldin A and an equimolar amount of DMSO as a background control. After 24 hours, blood was treated with EDTA (2 mM) for 15 minutes followed by red blood cell lysis and white cell fixation using FACS lysing solution (BD Biosciences) for 10 minutes. Cells were then cryopreserved in freezing media (90% fetal bovine serum (FBS) and 10% DMSO) and stored at –80°C until batched analysis.

### Cell staining and flow cytometry

Cell staining was performed on cryopreserved fixed cells that were thawed and washed with 1% FACS washing buffer (1% FBS in PBS). Cells were stained with the following surface antibody markers: CD4 ECD (SFCI12T4D11, Beckman Coulter), CD8 BV510 (RPA-8, Biolegend) and incubated at room temperature for 20 minutes. Cells were permeabilized and stained with intracellular antibody markers CD3 BV650 (OKT3), IFN-γ AlexaFluor® 700 (B27), TNF-α BV786 (Mab11) and IL-2 APC (MQ1-17H12) (all from Biolegend). Finally, cells were washed and fixed with Cellfix (BD Biosciences). Samples were acquired on a multiparameter BD Fortessa flow cytometer using Diva software version 8 and analyzed using FlowJo v10. Results are expressed as the frequency of CD4+ or CD8+ T cells expressing IFN-γ, TNF-α or IL-2. Cytokine responses presented are background subtracted values (from the frequency of cytokine produced by unstimulated cells).

## QUANTIFICATION AND STATISTICAL ANALYSIS

Statistical analyses were performed in Prism (v9.4.1; GraphPad Software Inc, San Diego, CA, USA). Non-parametric tests were used for all comparisons. The Mann-Whitney and Wilcoxon signed rank tests were used for unmatched and paired samples, respectively. Chi-square tests were used for comparisons between the proportion of responders represented as pie charts. All correlations reported are non-parametric Spearman’s correlations. Analysis of cytokine co-expressing populations was performed using SPICE version 5.1. *P* values less than 0.05 were considered statistically significant. Details of analyses performed for each experiment are described in the figure legends.

## Data Availability

All data produced in the present study are available upon reasonable request to the authors

## ACKNOWLEDGEMENTS

We thank the study participants and the parents/guardians of the children and the clinical staff and personnel at Red Cross War Memorial Children’s Hospital, the Drakenstein Child Health Study in Paarl, Western Cape and at Groote Schuur Hospital, Cape Town for their participation. We acknowledge Guarav Kwatra and Shabir Madhi for providing plasmids used for the endemic ELISA proteins. BEI resources provided the following reagents NIAID, NIH: Peptide Array, SARS-Related Coronavirus 2 Nucleocapsid (N) Protein, NR-52404 and SARS-Related Coronavirus 2 Membrane (M) Protein, NR-52403. This work is supported by the South African Medical Research Council (SA-MRC) with funds received from the South African Department of Science and Innovation (DSI), (grants 96825, SHIPNCD 76756, and DST/CON 0250/2012), the Poliomyelitis Research Foundation (21/65) and the Wellcome Centre for Infectious Diseases Research in Africa (CIDRI-Africa), which is supported by core funding from the Wellcome Trust (203135/Z/16/Z and 222754). SA-MRC to R.S.K. UK NIHR GECO award (GEC111 to H.J.Z), the Bill & Melinda Gates Foundation, USA (grants OPP1017641, OPP1017579 to H.J.Z). P.L.M. is supported by the South African Research Chairs Initiative of the Department of Science and Innovation and National Research Foundation of South Africa (NRF 9834), the SA Medical Research Council SHIP programme and the Centre for the AIDS Programme of Research in South Africa (CAPRISA). C.R. is supported by the EDCTP2 program of the European Union’s Horizon 2020 programme (TMA2017SF-1951-TB-SPEC), the Wellcome Trust (226137/Z/22/Z) and the National Institutes of Health (NIH) (R21AI148027). W.A.B. is supported by the EDCTP2 program of the European Union’s Horizon 2020 programme (TMA2016SF-1535-CaTCH-22), the Wellcome Trust (226137/Z/22/Z) and the EU-Africa Concerted Action on SARS-CoV-2 Virus Variant and Immunological Surveillance (COVICIS), funded through the EU’s Horizon Europe Research and Innovation Programme (101046041). H.J.Z is supported by the SA-MRC. LZ is funded by the South African Medical Research Council (SAMRC) through its Division of Research Capacity Development under the Mid-Career Scientist Programme from funding received from the South African National Treasury. The content hereof is the sole responsibility of the authors and do not necessarily represent the official views of the SAMRC. LJ Zühlke also receives support from the National Research Foundation of South Africa (NRFSA), as well as the UK Medical Research Council (MRC) and the UK Department for International Development (DFID) under the MRC/DFID Concordat agreement, via the African Research Leader Award (MR/S005242/1).

## AUTHOR CONTRIBUTIONS

R.S.K., K.W. and W.A.B. conceived the study and designed the experiments. N.S.B.B., A.N. and R.B. performed T cell assays. M.B.T., V.S. and A.W. performed SARS-CoV-2 ELISAs. S.V.G. and S.R.B. performed HCoV ELISAs. N.S.B.B., T.M.G., P.L.M., C.R., W.A.B. and R.S.K. analyzed the data. K.W., L.Z., and C.B. established and led the Red Cross childrens cohort. H.J.Z. established and led the Drakenstein Child Health cohort including a COVID sub-study. M.B and L.W. managed the Drakenstein Child Health cohort and repository. N.A.B.N. established and led the adult HCW cohort and M.M., S.S. and M.A. recruited participants and managed the cohort. N.S.B.B., C.R., W.A.B. and R.S.K. wrote the paper. All authors reviewed and edited the manuscript.

## REFERENCES

1. Aleebrahim-Dehkordi E, Soveyzi F, Deravi N, Rabbani Z, Saghazadeh A, Rezaei N. Human coronaviruses SARS-CoV, MERS-CoV, and SARS-CoV-2 in children. J Pediatr Nurs. 2021;56:70–79. doi:10.1016/j.pedn.2020.10.020

2. Chou J, Thomas PG, Randolph AG. Immunology of SARS-CoV-2 infection in children. Nat Immunol. 2022;23(2):177–185. doi:10.1038/s41590-021-01123-9

3. UNICEF. COVID-19 confirmed cases and deaths, age and sex-disaggregated data. Published April 2023. https://data.unicef.org/resources/covid-19-confirmed-cases-and-deaths-dashboard/. Accessed April 27, 2023.

4. Kim L, Whitaker M, Kambhampati A, et al. Hospitalization Rates and Characteristics of Children Aged <18 Years Hospitalized with Laboratory-Confirmed COVID-19 — COVID-NET, 14 States, March 1–July 25, 2020. 2020;69(32):1081–1088. https://www.cdc.gov/coronavirus/2019-ncov/covid-data/covidview/purpose-

5. COVID-NET. COVID-19-Associated Hospitalization Surveillance Network, Centers for Disease Control and Prevention. Published April 2023. https://gis.cdc.gov/grasp/covidnet/covid19_5.html. Accessed April 27, 2023

6. NICD. National Institute for Communicable Disease Monthly COVID-19 in Children.; 2022. https://www.nicd.ac.za/diseases-a-z-index/disease-index-covid-19/surveillance-reports/monthly-covid-19-in-children/. Accessed April 27, 2023

7. Nair H, Brooks A, Katz M, et al. Global burden of respiratory infections due to seasonal influenza in young children: a systematic review and meta-analysis. The Lancet. 2011;378:1917–1930. doi:10.1016/S0140

8. Bogunovic D, Merad M. Children and SARS-CoV-2. Cell Host Microbe. 2021;29(7):1040–1042. doi:10.1016/j.chom.2021.06.015

9. Williams N, Radia T, Harman K, Agrawal P, Cook J, Gupta A. COVID-19 severe acute respiratory syndrome coronavirus 2 (SARS-CoV-2) infection in children and adolescents: a systematic review of critically unwell children and the association with underlying comorbidities. Eur J Pediatr. 2021;180(10):3251–3252. doi:10.1007/s00431-021-04036-9

10. Tsankov BK, Allaire JM, Irvine MA, et al. Severe COVID-19 Infection and Pediatric Comorbidities: A Systematic Review and Meta-Analysis. International Journal of Infectious Diseases. 2021;103:246–256. doi:10.1016/j.ijid.2020.11.163

11. Sandoval M, Nguyen DT, Vahidy FS, Graviss EA. Risk factors for severity of COVID-19 in hospital patients age 18-29 years. PLoS One. 2021;16(7):1–22. doi:10.1371/journal.pone.0255544

12. Koch CM, Prigge AD, Anekalla KR, et al. Age-related Differences in the Nasal Mucosal Immune Response to SARS-CoV-2. Am J Respir Cell Mol Biol. 2022;66(2):206–222. doi:10.1165/rcmb.2021-0292OC

13. Muus C, Luecken MD, Eraslan G, et al. Single-cell meta-analysis of SARS-CoV-2 entry genes across tissues and demographics. Nat Med. 2021;27(3):546–559. doi:10.1038/s41591-020-01227-z

14. Schuler BA, Habermann AC, Plosa EJ, et al. Age-determined expression of priming protease TMPRSS2 and localization of SARS-CoV-2 in lung epithelium. J Clin Investig. 2021;131(1):1–7. doi:10.1172/JCI140766

15. Pierce CA, Sy S, Galen B, et al. Natural mucosal barriers and COVID-19 in children. JCI Insight. 2021;6(9):1–10. doi:10.1172/jci.insight.148694

16. Neeland MR, Bannister S, Clifford V, et al. Innate cell profiles during the acute and convalescent phase of SARS-CoV-2 infection in children. Nat Commun. 2021;12(1):1–5. doi:10.1038/s41467-021-21414-x

17. Speranza E. Children primed and ready for SARS-CoV-2. Nat Microbiol. 2021;6(11):1337–1338. doi:10.1038/s41564-021-00984-y

18. Loske J, Röhmel J, Lukassen S, et al. Pre-activated antiviral innate immunity in the upper airways controls early SARS-CoV-2 infection in children. Nat Biotechnol. 2022;40(3):319–324. doi:10.1038/s41587-021-01037-9

19. Yoshida M, Worlock KB, Huang N, et al. Local and systemic responses to SARS-CoV-2 infection in children and adults. Nature. 2021;602(7896):321–327. doi:10.1038/s41586-021-04345-x

20. Cotugno N, Ruggiero A, Bonfante F, et al. Virological and immunological features of SARS-CoV-2-infected children who develop neutralizing antibodies. Cell Rep. 2021;34(11):1–e5. doi:10.1016/j.celrep.2021.108852

21. Méndez-Echevarría A, Sainz T, Falces-Romero I, et al. Long-Term Persistence of Anti-SARS-CoV-2 Antibodies in a Pediatric Population. Pathogens. 2021;10(6):1–7. doi:10.3390/pathogens10060700

22. Dowell AC, Butler MS, Jinks E, et al. Children develop robust and sustained cross-reactive spike-specific immune responses to SARS-CoV-2 infection. Nat Immunol. 2022;23(1):40–49. doi:10.1038/s41590-021-01089-8

23. Cohen CA, Li APY, Hachim A, et al. SARS-CoV-2 specific T cell responses are lower in children and increase with age and time after infection. Nat Commun. 2021;12(1):1–14. doi:10.1038/s41467-021-24938-4

24. Humbert M, Olofsson A, Wullimann D, et al. Functional SARS-CoV-2 cross-reactive CD4 + T cells established in early childhood decline with age. Proc Natl Acad Sci. 2023;120(12):1–12. doi:10.1073/pnas.2220320120

25. Woudenberg T, Pelleau S, Anna F, et al. Humoral immunity to SARS-CoV-2 and seasonal coronaviruses in children and adults in north-eastern France. EBioMedicine. 2021;70:1–10. doi:10.1016/j.ebiom.2021.103495

26. Nickbakhsh S, Ho A, Marques DFP, McMenamin J, Gunson RN, Murcia PR. Epidemiology of Seasonal Coronaviruses: Establishing the Context for the Emergence of Coronavirus Disease 2019. J Infect Dis. 2020;222(1):17–25. doi:10.1093/infdis/jiaa185

27. Zimmermann C, Curtis N. Coronavirus Infections in Children Including COVID-19. Pediatr Infect Dis J. 2020;39(5):355–368. doi:10.1097/INF.0000000000002660

28. Fehr R A PS. Coronaviruses: An Overview of Their Replication and Pathogenesis. Methods Mol Biol. 2015;1282:1–23. doi:10.1007/978-1-4939-2438-7_1.

29. Park S, Lee Y, Michelow IC, Choe YJ. Global Seasonality of Human Coronaviruses: A Systematic Review. Open Forum Infect Dis. 2020;7(11):1–7. doi:10.1093/ofid/ofaa443

30. Audi A, AlIbrahim M, Kaddoura M, Hijazi G, Yassine HM, Zaraket H. Seasonality of Respiratory Viral Infections: Will COVID-19 Follow Suit? Front Public Health. 2020;8:1–8. doi:10.3389/fpubh.2020.567184

31. Sette A, Crotty S. Adaptive immunity to SARS-CoV-2 and COVID-19. Cell. 2021;184(4):861–880. doi:10.1016/j.cell.2021.01.007

32. Garrido C, Hurst JH, Lorang CG, et al. Asymptomatic or mild symptomatic SARS-CoV-2 infection elicits durable neutralizing antibody responses in children and adolescents. JCI Insight. 2021;6(17):1–13. doi:10.1172/jci.insight.150909

33. Dan JM, Mateus J, Kato Y, et al. Immunological memory to SARS-CoV-2 assessed for up to 8 months after infection. Science. 2021;371(6529):1–23. doi:10.1126/science.abf4063

34. Guo L, Wang G, Wang Y, et al. SARS-CoV-2-specific antibody and T-cell responses 1 year after infection in people recovered from COVID-19: a longitudinal cohort study. Lancet Microbe. 2022;3(5):e348–e356. doi:10.1016/S2666-5247(22)00036-2

35. Zuo J, Dowell AC, Pearce H, et al. Robust SARS-CoV-2-specific T cell immunity is maintained at 6 months following primary infection. Nat Immunol. 2021;22(5):620–626. doi:10.1038/s41590-021-00902-8

36. Pierce CA, Preston-Hurlburt P, Dai Y, et al. Immune responses to SARS-CoV-2 infection in hospitalized pediatric and adult patients. Sci Transl Med. 2020;12(564):1–11. doi:10.1126/scitranslmed.abe8120

37. Kaaijk P, Pimentel VO, Emmelot ME, et al. Children and Adults With Mild COVID-19: Dynamics of the Memory T Cell Response up to 10 Months. Front Immunol. 2022;13:1–15. doi:10.3389/fimmu.2022.817876

38. Tosif S, Neeland MR, Sutton P, et al. Immune responses to SARS-CoV-2 in three children of parents with symptomatic COVID-19. Nat Commun. 2020;11(1):1–8. doi:10.1038/s41467-020-19545-8

39. Tegally H, Wilkinson E, Giovanetti M, et al. Detection of a SARS-CoV-2 variant of concern in South Africa. Nature. 2021;592(7854):438–443. doi:10.1038/s41586-021-03402-9

40. Moss P. The T cell immune response against SARS-CoV-2. Nat Immunol. 2022;23(2):186–193. doi:10.1038/s41590-021-01122-w

41. Tsang HW, Chua GT, To KKW, et al. Assessment of SARS-CoV-2 Immunity in Convalescent Children and Adolescents. Front Immunol. 2021;12:1–10. doi:10.3389/fimmu.2021.797919

42. Paul K, Sibbertsen F, Weiskopf D, et al. Specific CD4+ T Cell Responses to Ancestral SARS-CoV-2 in Children Increase With Age and Show Cross-Reactivity to Beta Variant. Front Immunol. 2022;13:1–12. doi:10.3389/fimmu.2022.867577

43. Grifoni A, Weiskopf D, Ramirez SI, et al. Targets of T Cell Responses to SARS-CoV-2 Coronavirus in Humans with COVID-19 Disease and Unexposed Individuals. Cell. 2020;181(7):1489–1501. doi:10.1016/j.cell.2020.05.015

44. Mateus J, Grifoni A, Tarke A, et al. Selective and cross-reactive SARS-CoV-2 T cell epitopes in unexposed humans. Science. 2020;370(6512):89–94. doi:10.1126/science.abd3871

45. Le Bert N, Tan AT, Kunasegaran K, et al. SARS-CoV-2-specific T cell immunity in cases of COVID-19 and SARS, and uninfected controls. Nature. 2020;584(7821):457–462. doi:10.1038/s41586-020-2550-z

46. Swadling L, Diniz MO, Schmidt NM, et al. Pre-existing polymerase-specific T cells expand in abortive seronegative SARS-CoV-2. Nature. 2021;601(7891):110–117. doi:10.1038/s41586-021-04186-8

47. Tan HX, Lee WS, Wragg KM, et al. Adaptive immunity to human coronaviruses is widespread but low in magnitude. Clin Transl Immunology. 2021;10(3):1–15. doi:10.1002/cti2.1264

48. Singh V, Obregon-Perko V, Lapp SA, et al. Limited induction of SARS-CoV-2– specific T cell responses in children with multisystem inflammatory syndrome compared with COVID-19. JCI Insight. 2022;7(4):1–13. doi:10.1172/jci.insight.155145

49. Shah MM, Winn A, Dahl RM, Kniss KL, Silk BJ, Killerby ME. Seasonality of Common Human Coronaviruses, United States, 2014-2021. Emerg Infect Dis. 2022;28(10):1970–1976. doi:10.3201/eid2810.220396

50. de Vries RD. SARS-CoV-2-specific T-cells in unexposed humans: presence of cross-reactive memory cells does not equal protective immunity. Signal Transduct Target Ther. 2020;5(1):1–2. doi:10.1038/s41392-020-00338-w

51. Zar HJ, Nicol MP, MacGinty R, et al. Antibodies to seasonal coronaviruses rarely cross-react with SARS-CoV-2: findings from an African birth cohort. Pediatr Infect Dis J. 2021;40(12):e516–e519. doi:10.1097/INF.0000000000003325

52. Da Silva Antunes R, Pallikkuth S, Williams E, et al. Differential T-Cell Reactivity to Endemic Coronaviruses and SARS-CoV-2 in Community and Health Care Workers. J Infect Dis. 2021;224(1):70–80. doi:10.1093/infdis/jiab176

53. Woldemeskel BA, Kwaa AK, Garliss CC, Laeyendecker O, Ray SC, Blankson JN. Healthy donor T cell responses to common cold coronaviruses and SARS-CoV-2. J Clin Investig. 2020;130(12):6631–6638. doi:10.1172/JCI143120

54. Loyal L, Braun J, Henze L, et al. Cross-reactive CD4+ T cells enhance SARS-CoV-2 immune responses upon infection and vaccination. Science. 2021;374(6564):1–11. doi:10.1126/science.abh1823

55. Nelde A, Bilich T, Heitmann JS, et al. SARS-CoV-2-derived peptides define heterologous and COVID-19-induced T cell recognition. Nat Immunol. 2021;22(1):74–85. doi:10.1038/s41590-020-00808-x

56. Kundu R, Narean JS, Wang L, et al. Cross-reactive memory T cells associate with protection against SARS-CoV-2 infection in COVID-19 contacts. Nat Commun. 2022;13(1):1–8. doi:10.1038/s41467-021-27674-x

57. Yu ED, Narowski TM, Wang E, et al. Immunological memory to common cold coronaviruses assessed longitudinally over a three-year period pre-COVID19 pandemic. Cell Host Microbe. 2022;30(9):1269–1278. doi:10.1016/j.chom.2022.07.012

58. Saletti G, Gerlach T, Jansen JM, et al. Older adults lack SARS CoV-2 cross-reactive T lymphocytes directed to human coronaviruses OC43 and NL63. Sci Rep. 2020;10(1):1–10. doi:10.1038/s41598-020-78506-9

59. Niessl J, Sekine T, Lange J, et al. Identification of resident memory CD8 + T cells with functional specificity for SARS-CoV-2 in unexposed oropharyngeal lymphoid tissue. Sci Immunol. 2021;6(64):1–11. doi: 10.1126/sciimmunol.abk0894

60. Sagar M, Reifler K, Rossi M, et al. Recent endemic coronavirus infection is associated with less-severe COVID-19. J Clin Investig. 2021;131(1):1–5. doi:10.1172/JCI143380

61. Da Silva Antunes R, Pallikkuth S, Williams E, et al. Differential T-Cell Reactivity to Endemic Coronaviruses and SARS-CoV-2 in Community and Health Care Workers. J Infect Dis. 2021;224(1):70–80. doi:10.1093/infdis/jiab176

62. Dowell AC, Ireland G, Zuo J, Moss P, Ladhani S. Association of Spike-Specific T Cells with Relative Protection from Subsequent SARS-CoV-2 Omicron Infection in Young Children. JAMA Pediatr. 2022;177(1):96–97. doi:10.1001/jamapediatrics.2022.3868

63. Seder RA, Darrah PA, Roederer M. T-cell quality in memory and protection: Implications for vaccine design. Nat Rev Immunol. 2008;8(4):247–258. doi:10.1038/nri2274

64. Snyder LD, Chan C, Kwon D, et al. Polyfunctional T-cell signatures to predict protection from cytomegalovirus after lung transplantation. Am J Respir Crit Care Med. 2016;193(1):78–85. doi:10.1164/rccm.201504-0733OC

65. Ciuffreda D, Comte D, Cavassini M, et al. Polyfunctional HCV-specific T-cell responses are associated with effective control of HCV replication. Eur J Immunol. 2008;38(10):2665–2677. doi:10.1002/eji.200838336

66. Betts MR, Nason MC, West SM, et al. HIV nonprogressors preferentially maintain highly functional HIV-specific CD8+ T cells. Blood. 2006;107(12):4781–4789. doi:10.1182/blood-2005-12-4818

67. Altmann DM, Boyton RJ. SARS-CoV-2 T cell immunity: Specificity, function, durability, and role in protection. Sci Immunol. 2020;5(49):1–5. doi:10.1126/sciimmunol.abd6160

68. Bertoletti A, Le Bert N, Tan AT. SARS-CoV-2-specific T cells in the changing landscape of the COVID-19 pandemic. Immunity. 2022;55(10):1764–1778. doi:10.1016/j.immuni.2022.08.008

69. Spear TT, Evavold BD, Baker BM, Nishimura MI. Understanding TCR affinity, antigen specificity, and cross-reactivity to improve TCR gene-modified T cells for cancer immunotherapy. *Cancer Immunol*, Immunother. 2019;68(11):1881–1889. doi:10.1007/s00262-019-02401-0

70. Stone JD, Chervin AS, Kranz DM. T-cell receptor binding affinities and kinetics: impact on T-cell activity and specificity. Immunology. 2009;126(2):165–176. doi:10.1111/j.1365-2567.2008.03015.x

71. Tan MP, Gerry AB, Brewer JE, et al. T cell receptor binding affinity governs the functional profile of cancer-specific CD8+ T cells. Clin Exp Immunol. 2015;180(2):255–270. doi:10.1111/cei.12570

72. Jarjour NN, Masopust D, Jameson SC. T Cell Memory: Understanding COVID-19. Immunity. 2021;54(1):14–18. doi:10.1016/j.immuni.2020.12.009

73. Rydyznski Moderbacher C, Ramirez SI, Dan JM, et al. Antigen-Specific Adaptive Immunity to SARS-CoV-2 in Acute COVID-19 and Associations with Age and Disease Severity. Cell. 2020;183(4):996–1012.e19. doi:10.1016/j.cell.2020.09.038

74. Mahnke YD, Brodie TM, Sallusto F, Roederer M, Lugli E. The who’s who of T-cell differentiation: Human memory T-cell subsets. Eur J Immunol. 2013;43(11):2797–2809. doi:10.1002/eji.201343751

75. Ssemaganda A, Nguyen HM, Nuhu F, et al. Expansion of cytotoxic tissue-resident CD8+ T cells and CCR6+CD161+ CD4+ T cells in the nasal mucosa following mRNA COVID-19 vaccination. Nat Commun. 2022;13(1):1–9. doi:10.1038/s41467-022-30913-4

76. Lim JME, Tan AT, Bert N Le, Hang SK, Low JGH, Bertoletti A. SARS-CoV-2 breakthrough infection in vaccinees induces virus-specific nasal-resident CD8+ and CD4+ T cells of broad specificity. J Exp Med. 2022;219(10):1–10. doi:10.1084/jem.20220780

77. Weatherhead JE, Clark E, Vogel TP, Atmar RL, Kulkarni PA. Inflammatory syndromes associated with SARS-cov-2 infection: Dysregulation of the immune response across the age spectrum. J Clin Investig. 2020;130(12):6194–6197. doi:10.1172/JCI145301

78. Zar HJ, Barnett W, Myer L, Stein DJ, Nicol MP. Investigating the early-life determinants of illness in Africa: the Drakenstein Child Health Study. Thorax. 2015;70(6):592–594. doi:10.1136/thoraxjnl-2014-206242

79. Zar HJ, MacGinty R, Workman L, et al. Natural and hybrid immunity following four COVID-19 waves: A prospective cohort study of mothers in South Africa. EClinicalMedicine. 2022;53:1–12. doi:10.1016/j.eclinm.2022.101655

80. Makatsa MS, Tincho MB, Wendoh JM, et al. SARS-CoV-2 Antigens Expressed in Plants Detect Antibody Responses in COVID-19 Patients. Front Plant Sci. 2021;12:1–13. doi:10.3389/fpls.2021.589940

81. Mutavhatsindi H, Riou C. Protocol to quantify and phenotype SARS-CoV-2-specific T cell response using a rapid flow-cytometry-based whole blood assay. STAR Protoc. 2022;3(4):1–15. doi:10.1016/j.xpro.2022.101771

82. Riou C, Schäfer G, du Bruyn E, et al. Rapid, simplified whole blood-based multiparameter assay to quantify and phenotype SARS-CoV-2-specific T-cells. Eur Respir. 2022;59(1):1–13. doi:10.1183/13993003.00285-2021

