## Supplemental Data for "Distinct T cell functional profiles in SARS-CoV-2 seropositive and seronegative children associated with endemic human coronavirus cross-reactivity"

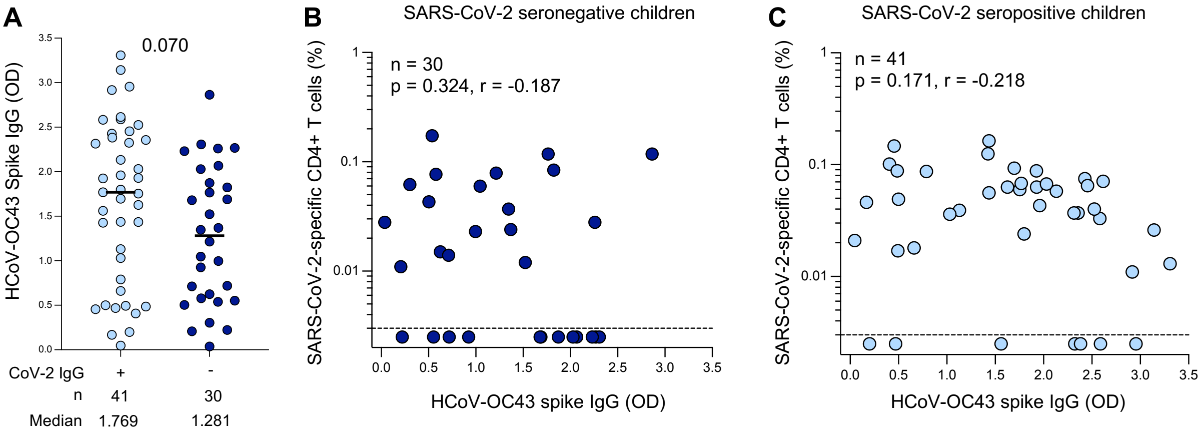


**Supplemental Figure 1: SARS-CoV-2 cross-reactivity to endemic HCoV-OC43 in children**

(A) The magnitude of HCoV-OC43 spike IgG levels were measured by ELISA in SARS-CoV-2 seropositive (light blue; n=41) and seronegative (dark blue; n=30) children. The bars represent the median values. A statistical comparison was performed using the Mann-Whitney test between seropositive and seronegative children; a p value <0.05 was considered statistically significant.

(B) Correlation between the frequency of SARS-CoV-2-specific CD4+ T cells and HCoV-OC43-spike IgG levels in SARS-CoV-2 seronegative children (n=30).

(C) Correlation between the frequency of SARS-CoV-2-specific CD4+ T cells and HCoV-OC43-spike IgG levels in SARS-CoV-2 seropositive children (n=41). Statistical comparisons in (B) and (C) were performed using a two-tailed non-parametric Spearman rank tests; p values <0.05 were considered statistically significant and are bolded and correlation coefficients are shown.
